# Feasibility and Safety of Perioperative Chemotherapy with Fluorouracil plus Leucovorin, Oxaliplatin, and Docetaxel (FLOT) for Locally-Advanced Gastric Cancer Patients in China

**DOI:** 10.1101/2020.05.22.20110668

**Authors:** Sah Birendra Kumar, Xu Wei, Zhang Benyan, Zhang Huan, Yuan Fei, Li Jian, Liu Wentao, Chao Yan, Li Chen, Yan Min, Zhu Zhenggang

## Abstract

**Background:** Neoadjuvant fluorouracil plus leucovorin, oxaliplatin, and docetaxel (FLOT) has shown significant benefits for gastric cancer patients. However, it has not been well accepted in Asian countries. We conducted a prospective study on the safety and feasibility of the FLOT regimen in Chinese patients.

**Methods:** Patients with adenocarcinoma of the stomach or esophagogastric junction received 4 cycles of neoadjuvant chemotherapy (NAC) and 4 cycles of adjuvant chemotherapy (AC) with the FLOT regimen. The completion status of chemotherapy, adverse events, postoperative morbidities, and pathological tumor regression were analyzed. The two-year overall survival (OS) and relapse-free survival are presented.

**Results:** Altogether, 10 patients were enrolled, and all patients completed 4 cycles of neoadjuvant chemotherapy. There were no severe hematological adverse events (grade 3 or above), except for a case of grade 3 anemia. All 10 patients underwent radical gastrectomy. Nine patients had R0 resection, and 3 patients had complete or subtotal pathological tumor regression. Nine patients completed 4 cycles of adjuvant chemotherapy, but only one patient completed the full dose of adjuvant chemotherapy. The dose of adjuvant chemotherapy was reduced by 25% or less in the other patients. The median follow-up time was 23.13 months, 8 patients achieved the overall survival endpoint, and 7 patients had relapse-free survival for this period. Two patients died of disease progression.

**Conclusions:** Our study demonstrates that the neoadjuvant FLOT regimen is safe and effective for Chinese patients. Dose adjustment is necessary for adjuvant chemotherapy. The pathological regression and survival rates need reevaluation in a larger cohort.

The trial is registered with ClinicalTrials.gov (number NCT03646591).

## Background

Neoadjuvant chemotherapy (NAC) for gastric cancer gained attention after the publication of the MAGIC trial in the New England Journal of Medicine in 2006, which advocated for perioperative chemotherapy with triplet chemotherapy consisting of epirubicin, cisplatin, and infusional fluorouracil (1). However, NAC has been debated for a long time among various centers around the world. It was interesting that most of the chemotherapy- or chemoradiotherapy-related trials came from Western countries despite the higher prevalence of gastric cancer in Eastern countries(1, 2). Japanese researchers conducted most of the trials for the last few decades, but there have been no large-scale phase 3 trials on NAC for resectable locally advanced gastric cancer (LAGC) patients, and NAC was only tested in Japan for gastric cancer patients with bulky metastatic lymph nodes, para-aortic lymph node (PAN)-positive patients, or those with large ulcero-invasive type (Bormann type 3) or linitis plastic (Borrmann type 4) gastric cancer(3-6). Even if NAC was safe in Japanese patients, its effect on survival was not clearly justified (7, 8). In addition, the chemotherapy regimens were relatively conservative in Japan compared to the chemotherapy regimens used in Western countries, and these regimens were mainly based on cisplatin and fluorouracil or S1(3-6, 9). The addition of taxane-based chemotherapy failed to show benefits for gastric cancer patients in Japan(9). Adjuvant chemotherapy was suggested to be beneficial for gastric cancer patients, and a review was long before published on JAMA by French researchers(10). In general, for patients with resectable LAGC, standard gastrectomy with adjuvant chemotherapy is well accepted in Asian countries, especially after the publication of the milestone articles of the CLASSIC and ARTIST trials from South Korea(11-13). Adjuvant chemotherapy was mainly dominated by platinum-based doublet chemotherapy. Oral capecitabine and cisplatin or oxaliplatin are common adjuvant chemotherapy agents in Eastern countries (11, 13). On the other hand, German scientists published a series of clinical studies on neoadjuvant chemotherapy with taxane-based chemotherapy (14-16). The German studies even advocated for taxane-based triplet chemotherapy(16, 17).The groundbreaking results comparing ECF or ECX with FLOT showed that FLOT was associated with significantly higher proportions of patients achieving pathological complete regression than ECF/ECX(17). NAC with taxane-based chemotherapy was later supported by Italian researchers (18, 19).

The FLOT regimen is a taxane-based triplet chemotherapy regimen that is generally considered more toxic. Zhou et al. investigated and recommended a modified FLOT regimen; however, this study lacks the evidence to support this modified regimen (20). Furthermore, initial studies of FLOT, which were also published by Lancet, showed that it was well tolerated in gastric cancer patients with promising results in terms of pathological regression and survival in Germany(16, 17).

There have been many published articles including those in high impact major journals which suggest that the FLOT regimen is safe and tolerable(16,17). The main purpose of this study was to explore whether the standard dose of the FLOT regimen is also tolerable in Chinese patients. Therefore, there is a scientific basis for conducting a prospective study to evaluate the safety and feasibility of a standard FLOT regimen for Chinese patients rather than conducting a dose-finding study. In this cohort, we strictly adhered to the FLOT regimen as described by the original investigators (16).

## Methods

This is an open-label, single-arm prospective study. We assessed the FLOT regimen for safety and feasibility in Chinese gastric cancer patients. Patients were enrolled between November 2017 and August 2018 from Ruijin Hospital, Shanghai Jiaotong University School of Medicine. The trial is registered with ClinicalTrials.gov, number NCT03646591.

### Inclusion criteria

Age: 18-80 years old.

Sex: all.

Diagnosis: histologically-confirmed adenocarcinoma of the stomach or esophagogastric junction.

Clinical stage: stage III or above (CT3-4bN1-3M0, AJCC/UICC 8^th^ cTNM staging system).

Performance status: Eastern Cooperative Oncology Group (ECOG) ≤ 2 (normal to symptomatic but in bed less than half the day).

Clinically fit for systemic chemotherapy and gastric cancer surgery, i.e., adequate renal, hepatic, hematologic, and pulmonary function.

Written informed consent provided.

### Exclusion criteria

Clinically unfit for systemic chemotherapy and gastric cancer surgery, i.e. uncontrolled cardiac disease or other clinically significant uncontrolled comorbidities.

Unable to undergo general anesthesia

Distant metastases (including peritoneal or retroperitoneal lymph node metastases)

Locally advanced inoperable disease (clinical assessment)

Relapse of gastric cancer.

Second malignant disease

Prior chemotherapy or radiotherapy

Inclusion in another clinical trial

Known contraindications or hypersensitivity to planned chemotherapy

### Pretreatment assessment

All patients underwent a full clinical assessment before the commencement of the treatment, which included a full medical history, physical examination, complete blood count, clotting analysis, serum liver function, renal function test, 24-hour urinary clearance, and blood tumor markers for gastrointestinal diseases. Electrocardiography, echocardiography, chest radiography, and computed tomography of the chest, abdomen and pelvis, and upper gastrointestinal endoscopy were performed. The specially designed protocol was used for staging the CT scans of gastric cancer, consisting of arterial, venous, and portal phase transverse section images and reconstruction images of the sagittal and coronary sections. Ultrasonography and magnetic resonance (MR) were used if clinically necessary to rule out suspicious distant metastases or retroperitoneal lymph nodes. Positron emission tomography (PET) or whole-body bone scintigraphy was required in suspected cases. Diagnostic laparoscopy was performed to rule out peritoneal metastases in suspicious findings on CT. In this cohort, the patients were only enrolled if they met all the inclusion criteria and we only collected the data of patients who were enrolled in this study. Patients with distant metastases including peritoneal metastases and retroperitoneal lymph node metastases were excluded according to exclusion criteria thus we don’t have data from other patients.

Clinical staging was performed according to the AJCC/UICC tumor-node-metastasis (TNM) eighth edition staging system.

### Neoadjuvant Chemotherapy

A standard dose of FLOT chemotherapy was prescribed (16). Preventive antiemetic and dexamethasone were allowed before chemotherapy, and growth factor or other supportive medicines were allowed for treatment only.

#### FLOT chemotherapy regimen

A cycle consists of the following:

Day 1: 5-FU 2600 mg/m^2^ intravenously

via peripherally inserted central catheter (PICC) for 24 hours

Day 1: Leucovorin 200 mg/m^2^ intravenously

Day 1: Oxaliplatin 85 mg/m^2^ intravenously

Day 1: Docetaxel 50 mg/m^2^ intravenously

Repeated every 15th day

### Dose alteration or stop

The National Cancer Institute Common Terminology Criteria for Adverse Events (CTCAE, 4.0) was followed for the evaluation of adverse effects. The postponement or dose adjustment of treatment was allowed after discussion with oncologists and was carefully documented. Decreasing of drug dose was allowed between 10 to 25 percent of the standard dose for the patients with grade 4 or above hematological or grade 3 or above non-hematological adverse events. Chemotherapy would be delayed or stopped for those patients who had refractory adverse events after dose adjustment for two times.

### Restaging

Two specialized radiologists independently evaluated the overall response rate. Any conflicting results were settled after discussion among both radiologists and investigators. The response to treatment was evaluated according to the Response Evaluation Criteria in Solid Tumors (RECIST, version 1.1) guidelines(21).

### Surgery

Surgery was scheduled between two to four weeks after the completion of the planned chemotherapy. The patients underwent an exploratory laparoscopic examination to rule out peritoneal or distant metastases. Surgery was terminated if there was peritoneal or distant metastasis. Standard gastrectomy with curative intent was the principal surgical procedure. It involves resection of at least two-thirds of the stomach with D2 lymph node dissection.

### Pathological assessment

Pathologists carefully examined residual vital tumor cells and the remnant of the previous tumor as necrosis, fibrosis, or scar. The tumor regression grading (TRG) with the Becker criteria was used for the evaluation of pathological response in the resected specimens(22). Two specialized pathologists independently rated the TRG grading.

### Endpoints

#### Primary Outcome Measure

Completion rate of the preoperative FLOT regimen

#### Secondary Outcome Measures

##### Adverse events

Pathological response rate: According to tumor regression grading (TRG)

Postoperative morbidity: Postoperative complications

Postoperative mortality: Death due to surgical complications

Overall survival (OS): Time from randomization to death from any cause

Relapse-free survival(RFS): Time from randomization to relapse

##### Sample size calculation

The sample size was not calculated by any statistical methods and it was estimated empirically. This research was intended to obtain preliminary data for Chinese patients and to pave the way for conducting further Phase II or III study.

### Statistical analysis

The statistical analysis was performed with Statistical Package for Social Science (SPSS) version 22.0 for Windows (SPSS, Inc., Chicago, Illinois). The continuous data(Age, BMI, interval time between two chemotherapy cycles) are expressed as the median and range. The number of cases was provided for categorical variables. Survival data were presented as the length of overall survival(OS), relapse-free survival(RFS) in months. A Kaplan-Meier plot was created for survival analysis.

## Results

Altogether, 10 patients were enrolled in this study (Table 1). All patients completed 4 cycles of FLOT chemotherapy before curative gastrectomy. The median time interval between two cycles of neoadjuvant chemotherapy was 15 days for all three intervals. Eight patients received a full dose of the standard preoperative FLOT chemotherapy regimen. The chemotherapy dose was reduced by 25 percent or less in two patients (Table 2). All 10 patients underwent surgery at the same hospital. One patient refused adjuvant chemotherapy. Nine patients completed 4 cycles of adjuvant chemotherapy. Eight patients received adjuvant chemotherapy at the same hospital, and one patient received adjuvant chemotherapy at a different hospital. The median time to the first chemotherapy cycle after surgery was 36 days. The median time was approximately 21 days between two chemotherapies. There were no severe hematological adverse events (grade 3 or above), except for a case with grade 3 anemia. Four patients had grade 3 or 4 vomiting, and all other non-hematological adverse events were grade 2 or below (Table 3).

**Table 1.** Demographic data

| Parameter |  | Number |
| --- | --- | --- |
| Sex | Male | 7 |
|  | Female | 3 |
| Age (years) | Median | 59.50 |
|  | Range | 26(43-69) |
| Body mass index | Median | 22.95 |
|  | Range | 17.29(16.30-33.59) |
| Site of tumor | Body | 4 |
|  | Distal | 6 |
| Type of resection | Partial | 7 |
|  | Total | 3 |
| Prechemotherapy time interval (days) | 1<sup>st</sup>-2<sup>nd</sup> chemo | 15(14-18) |
| Median (range) | 2<sup>nd</sup>-3<sup>rd</sup> chemo | 15(14-20) |
|  | 3<sup>rd</sup>-4<sup>th</sup> chemo | 15(11-17) |
| Postchemotherapy time interval (days) | Surgery-1<sup>st</sup> chemo | 36(26-50) |
| Median (range) | 1<sup>st</sup>-2<sup>nd</sup> chemo | 22(14-46) |
|  | 2<sup>nd</sup>-3<sup>rd</sup> chemo | 21(14-28) |
|  | 3<sup>rd</sup>-4<sup>th</sup> chemo | 22(14-40) |

**Table 2.** Preoperative chemotherapy dose alteration

| Case | 1<sup>st</sup> Chemo | 2<sup>nd</sup> Chemo | 3<sup>rd</sup> Chemo | 4<sup>th</sup> Chemo |
| --- | --- | --- | --- | --- |
| 1 | FLOT: Full | FLOT: Full | FLOT: Full | FLOT: Full |
| 2 | FLOT: Full | FLOT: Full | FLOT: Full | FLOT: Full |
| 3 | FLOT: Full | FLOT: Full | FLOT: Full | FLOT: Full |
| 4 | F: DRB 10-25% | F: DRB 10-25% | F: DRB 10-25% | F: DRB 10-25% |
|  | L: Full | L: Full | L: Full | L: Full |
|  | O: DRB 25% | O: DRB 25% | O: DRB 25% | O: DRB 25% |
|  | T: DRB 25% | T: DRB 25% | T: DRB 25% | T: DRB 25% |
| 5 | FLOT: Full | FLOT: Full | FLOT: Full | FLOT: Full |
| 6 | FLOT: Full | FLOT: Full | FLOT: Full | FLOT: Full |
| 7 | F: DRB 10-25% | F: DRB 10-25% | F: DRB 10-25% | F: DRB 10-25% |
|  | L: Full | L: Full | L: Full | L: Full |
|  | O: DRB 10-25% | O: DRB 10-25% | O: DRB 10-25% | O: DRB 10-25% |
|  | T: Full | T: Full | T: Full | T: Full |
| 8 | FLOT: Full | FLOT: Full | FLOT: Full | FLOT: Full |
| 9 | FLOT: Full | FLOT: Full | FLOT: Full | FLOT: Full |
| 10 | FLOT: Full | FLOT: Full | F: DRB 10-25% | F: DRB 10-25% |
|  |  |  | L: Full | L: Full |
|  |  |  | O: DRB 10-25% | O: DRB 10-25% |
|  |  |  | T: DRB 10-25% | T: DRB 10-25% |

DRB: Dose reduced by

**Table 3.** Adverse effects

| Parameter |  | Number |
| --- | --- | --- |
| WBC decreased | Grade 0 | 5 |
|  | Grade 1 | 4 |
|  | Grade 2 | 1 |
| Neutrophil count decreased | Grade 0 | 4 |
|  | Grade 1 | 1 |
|  | Grade 2 | 5 |
| Febrile neutropenia | Grade 0 | 10 |
| Anemia | Grade 0 | 2 |
|  | Grade 1 | 4 |
|  | Grade 2 | 3 |
|  | Grade 3 | 1 |
| Platelet count decreased | Grade 0 | 10 |
| AST increased | Grade 0 | 4 |
|  | Grade 1 | 6 |
| ALT increased | Grade 0 | 7 |
|  | Grade 1 | 3 |
| Nausea | Grade 0 | 5 |
|  | Grade 1 | 5 |
| Vomiting | Grade 0 | 6 |
|  | Grade 3,4 | 4 |
| Diarrhea | Grade 0 | 10 |
| Peripheral neuropathy | Grade 0 | 10 |
| Fatigue | Grade 0 | 10 |
| Anorexia | Grade 0 | 9 |
|  | Grade 2 | 1 |
| Oral mucositis | Grade 0 | 9 |
|  | Grade 1 | 1 |

### CT results

The prechemotherapy clinical TNM staging showed that all patients had advanced-stage tumors. Four patients had a partial response, and six patients had stable disease comparing the prechemotherapy and postchemotherapy radiological results according to the RECIST 1.1 criteria (Table 4).

**Table 4.** Computed tomography (CT) results

| Parameter |  | Number |
| --- | --- | --- |
| Prechemotherapy tumor(T) | T4A | 7 |
|  | T4B | 3 |
| Prechemotherapy lymph node(N) | N1 | 5 |
|  | N2 | 5 |
| Postchemotherapy tumor(T) | T3 | 2 |
|  | T4A | 7 |
|  | T4B | 1 |
| Postchemotherapy lymph node(N) | N0 | 2 |
|  | N1 | 5 |
|  | N2 | 3 |
| Response | Partial response(PR) | 4 |
|  | Stable disease(SD) | 6 |

### Postoperative complications

Standard curative gastrectomy (gastrectomy+ D2 lymphadenectomy) was performed in all 10 patients. Among them, 3 patients underwent total gastrectomy, and 7 patients underwent distal gastrectomy. The median postoperative hospital stay was 9 days, and five patients had minor or moderate scale postoperative complications. There were no cases of anastomotic leakage, reoperation, or death due to surgical complications (Table 5).

**Table 5.**
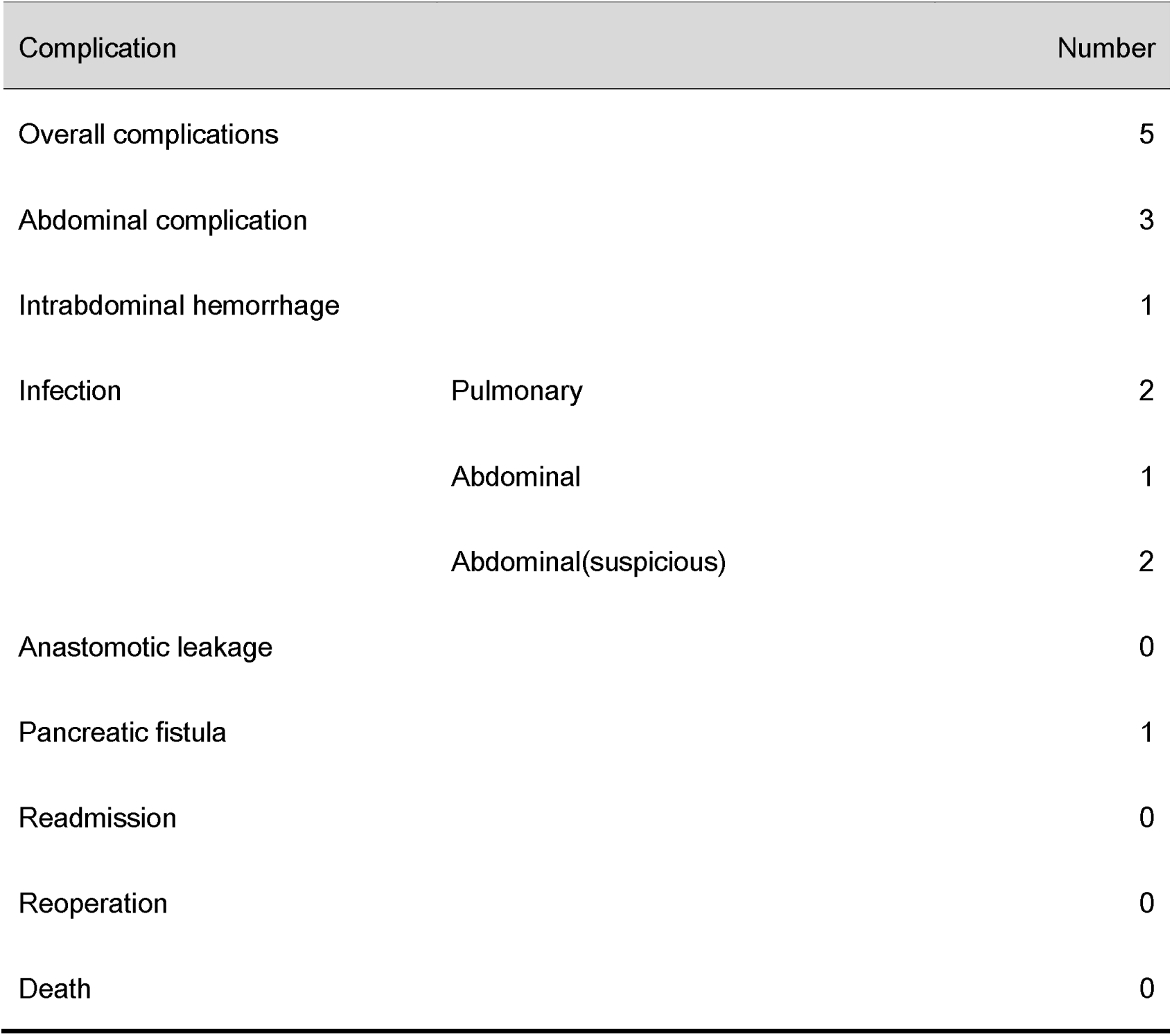
Postoperative complications

### Postoperative pathology

Pathological reports confirmed that 9 of 10 patients achieved R0 resection. One patient had tumor positive on the lower margin of the resected specimen, thus considered R1 resection. The median number of examined lymph nodes was 28, and eight patients were classified as stage III according to the ypTNM classification. One patient had complete tumor regression (TRG 1a), and two patients had subtotal tumor regression (Table 6).

**Table 6.**
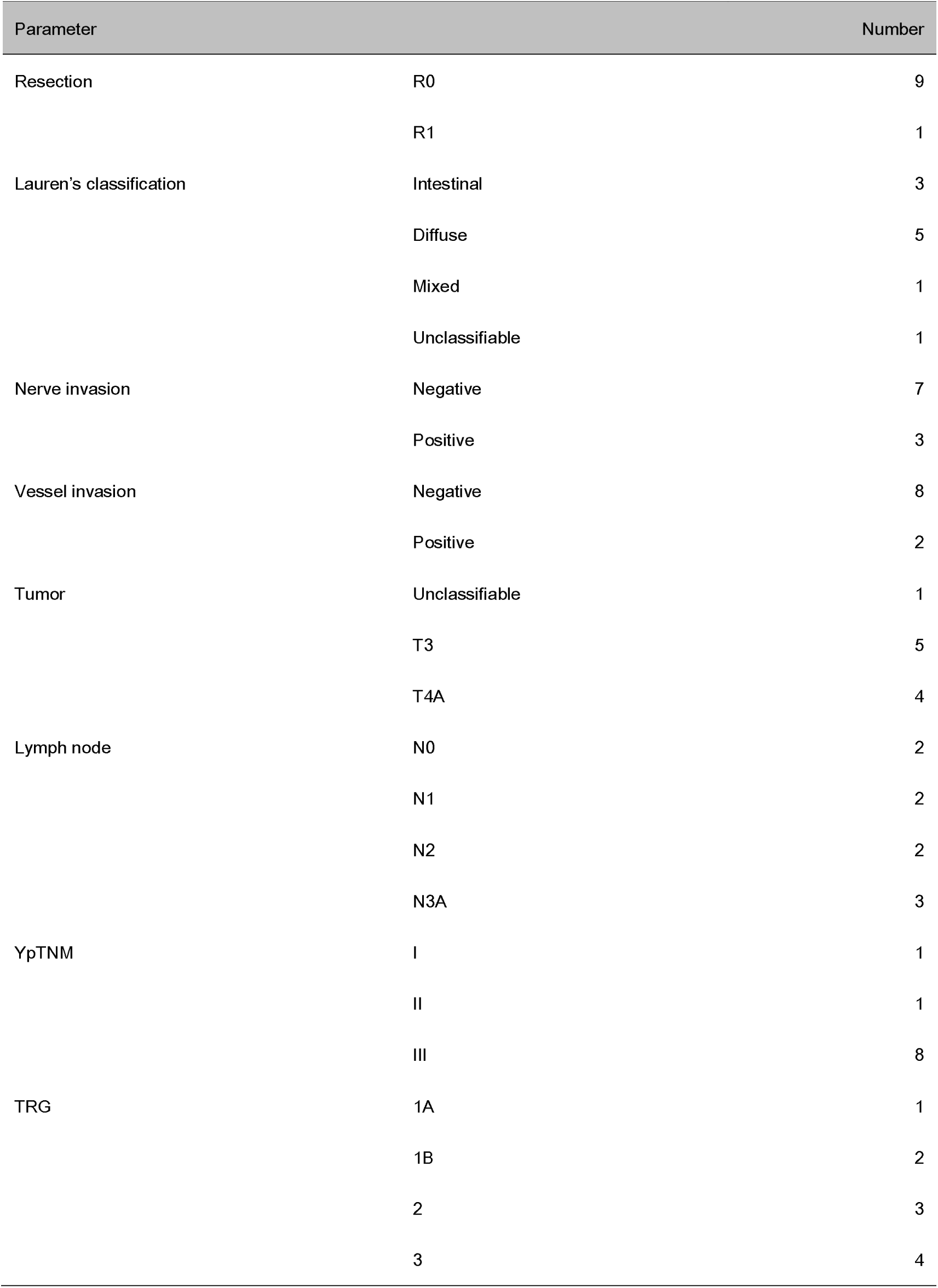
Postoperative pathology

| Parameter |  | Number |
| --- | --- | --- |
| Resection | R0 | 9 |
|  | R1 | 1 |
| Lauren's classification | Intestinal | 3 |
|  | Diffuse | 5 |
|  | Mixed | 1 |
|  | Unclassifiable | 1 |
| Nerve invasion | Negative | 7 |
|  | Positive | 3 |
| Vessel invasion | Negative | 8 |
|  | Positive | 2 |
| Tumor | Unclassifiable | 1 |
|  | T3 | 5 |
|  | T4A | 4 |
| Lymph node | N0 | 2 |
|  | N1 | 2 |
|  | N2 | 2 |
|  | N3A | 3 |
| YpTNM | I | 1 |
|  | II | 1 |
|  | III | 8 |
| TRG | 1A | 1 |
|  | 1B | 2 |
|  | 2 | 3 |
|  | 3 | 4 |

### Adjuvant chemotherapy

Nine patients completed 4 cycles of postoperative chemotherapy. One patient refused adjuvant chemotherapy. Only one patient completed the full dose of all four cycles of postoperative chemotherapy. The dose of adjuvant chemotherapy was reduced by 25% or less in the other patients (Table 7).

**Table 7.** Postoperative chemotherapy dose alteration

| Case | 1<sup>st</sup> Chemo | 2<sup>nd</sup>Chemo | 3<sup>rd</sup> Chemo | 4<sup>th</sup> Chemo |
| --- | --- | --- | --- | --- |
| 1 | FLOT: Full | FLOT: Full | FLOT: Full | FLOT: Full |
| 2 | FLOT: CAC | CAC | CAC | CAC |
| 3 | FLOT: DRB 10-25% | FLOT: DRB 10-25% | FLOT: DRB 10-25% | FLOT: DRB 10-25% |
| 4 | No chemotherapy | No chemotherapy | No chemotherapy | No chemotherapy |
| 5 | FLOT: DRB 10-25% | FLOT: DRB 10-25% | FLOT: DRB 10-25% | FLOT: DRB 10-25% |
| 6 | FLOT: DRB 10-25% | F: DRB 10-25% | F: DRB 10-25% | F: DRB 10-25% |
|  |  | L: Full | L: Full | L: Full |
|  |  | O: DRB 10-25% | O: DRB 10-25% | O: Full |
|  |  | T: Full | T: Full | T: Full |
| 7 | F: DRB 10-25% | F: DRB 10-25% | F: DRB 10-25% | F: DRB 10-25% |
|  | L: Full | L: Full | L: Full | L: Full |
|  | O: DRB 10-25% | O: DRB 10-25% | O: DRB 10-25% | O: DRB 10-25% |
|  | T: Full | T: Full | T: Full | T: Full |
| 8 | FLOT: DRB 10-25% | FLOT: DRB 10-25% | FLOT: Full | FLOT: DRB 10-25% |
| 9 | F: DRB 10-25% | FLOT: DRB 25%. | CAC | CAC |
|  | L: DRB 10-25% |  |  |  |
|  | O: DRB 25% |  |  |  |
|  | T: DRB 10-25% |  |  |  |
| 10 | FLOT: DRB 25% | FLOT: DRB 25% | FLOT: DRB 25% | FLOT: DRB 25% |

DRB: Dose reduced by, CAC: Chemotherapy at another center

### Follow up

All 10 patients had a timely follow-up, 8 patients were still alive; among them, 7 patients were without any signs of relapse at the last follow-up visit(14^th^ May 2020). One patient had a relapse of peritoneal and ovary metastases after eight months of the first chemotherapy cycle and was still undergoing chemotherapy treatment. Thus, 8 patients achieved overall survival (OS), and 7 patients achieved relapse-free survival (RFS) at the median follow up time of 23.13 months. Two patients died, one of whom died of retroperitoneal metastases. The OS time was 26 months, and the RFS time was 6 months for this patient. Another patient died of peritoneal metastases and Krukenberg tumors. The OS time was 10.6 months, and the RFS time was approximately 6 months for this patient (Table 8). Kaplan-Meier plot for OS and RFS are provided in Fig.1A and 1B.

**Fig. 1.**
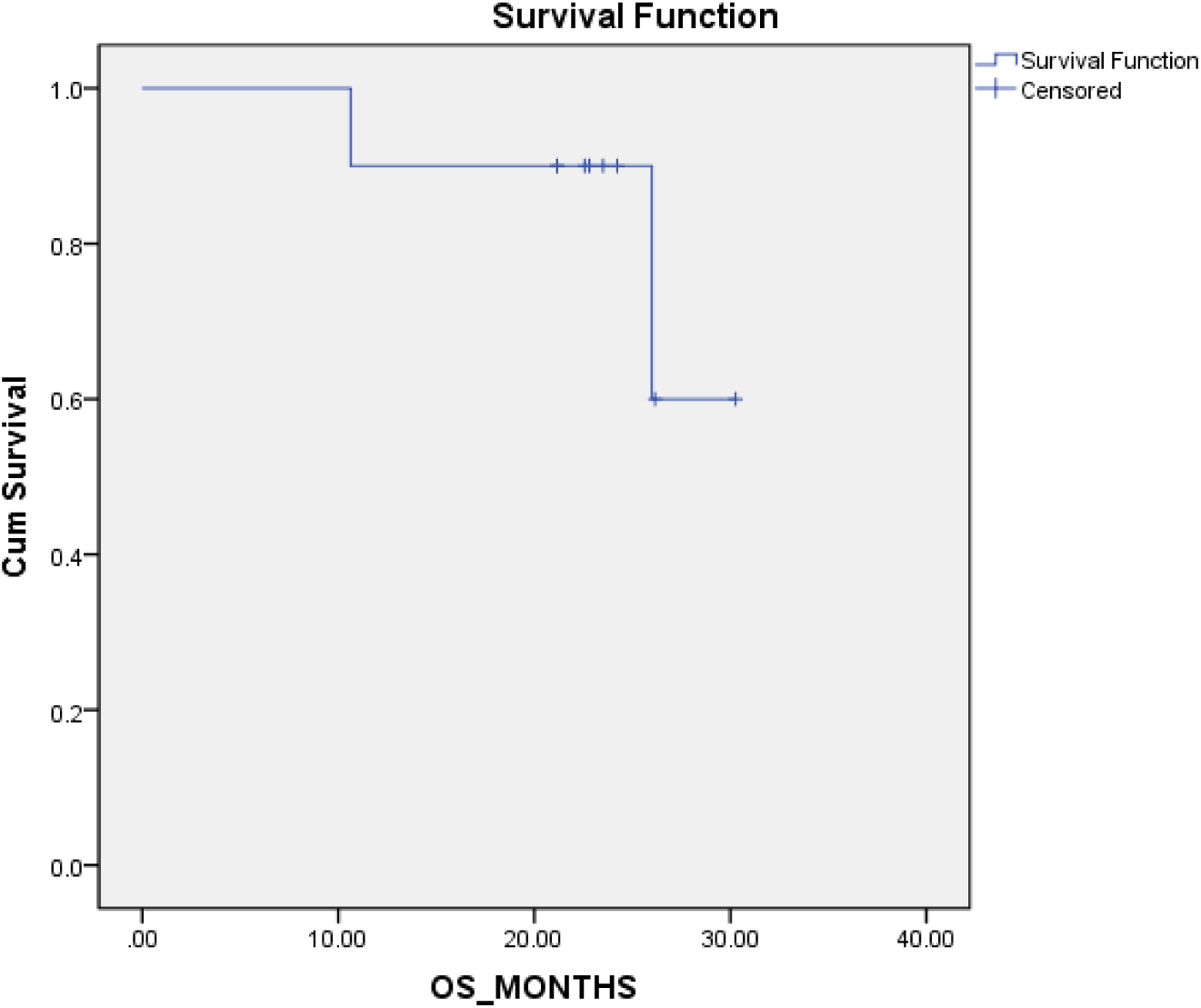
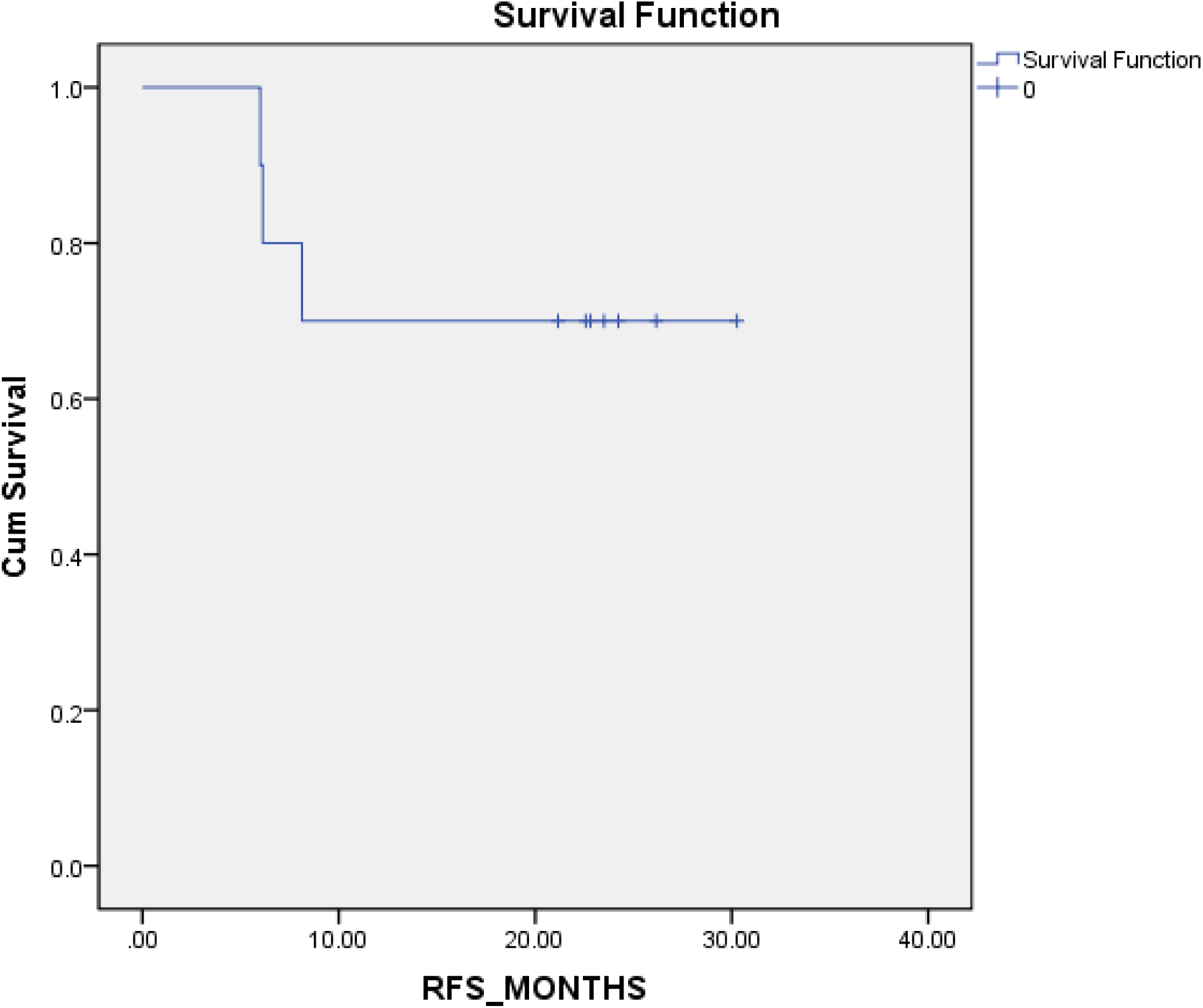
Kaplan-Meier(K-M) Plot for OS and RFS 1A K-M plot for OS 1B K-M plot for RFS

**Table 8.** Survival analysis

| Case | Status | Cause | RFS(days) | OS(days) |
| --- | --- | --- | --- | --- |
| 1 | Dead/Relapse | Retroperitoneal metastases | 180 | 780 |
| 2 | Alive | No sign of relapse | 727 | 727 |
| 3 | Alive | No sign of relapse | 705 | 705 |
| 4 | Alive | No sign of relapse | 677 | 677 |
| 5 | Dead/Relapse | Peritoneal metastases/Ovary | 184 | 319 |
| 6 | Alive | No sign of relapse | 684 | 684 |
| 7 | Alive | No sign of relapse | 908 | 908 |
| 8 | Alive | No sign of relapse | 635 | 635 |
| 9 | Alive | No sign of relapse | 785 | 785 |
| 10 | Alive/Relapse | Peritoneal metastases/Ovary | 244 | 635 |

## Discussion

Despite many presumptions about the toxic adverse effects, our study shows that the FLOT regimen is safe and effective in Chinese patients, as most of the patients tolerated the standard dose of the FLOT regimen, which may be due to the well-balanced dose combination of this regimen, which is different from other combinations(9, 16). All 4 cycles with a full dose of neoadjuvant chemotherapy were completed in 8 patients. Even for 2 patients, for whom the dose was adjusted, it was solely the decision of a clinical doctor; there was no concrete rationale for reducing the dose in those two cases. According to the study design, dose reduction was allowed between 10 to 25 percent. This is a cohort of 10 patients, for the consistency in data and easy understanding we recorded them in this range. Most of the hematological adverse effects and symptomatic adverse effects were acceptable for triplet chemotherapy. None of the patients discontinued chemotherapy due to adverse events.

The postoperative morbidities were acceptable for radical gastrectomy compared to previously reported results(23, 24). There were no deaths, anastomotic leakages, or reoperations due to postoperative complications, which are the main concerns of surgeons, especially for patients who received neoadjuvant chemotherapy.

The postoperative pathological findings confirmed that the majority of patients had advanced-stage adenocarcinoma; eight patients were pathologically confirmed as stage III in ypTNM staging. In previewing the prechemotherapy clinical stage, we found that all the tumors were T4a or T4b, and lymph nodes were positive on CT diagnosis. After chemotherapy, restaging showed that there was downstaging in four cases by the RECIST criteria. There was no progressive disease. This finding indicates that all patients achieved disease control. The pathological results showed that 9 of 10 patients achieved R0 resection, and pathological tumor regression was observed in 6 patients, including one patient who had a complete response. The TRG results were comparable with those reported in FLOT4(16); however, this is a relatively small number of patients, and this result needs to be re-evaluated in a larger cohort.

The completion rate of full-dose neoadjuvant chemotherapy was comparable with that in the FLOT4 study (16), but the only concern was the feasibility of the full dose of adjuvant chemotherapy, as the data showed that most of the patients were administered a reduced dose of the FLOT regimen despite completing all four adjuvant chemotherapy cycles. Even in the FLOT4 study, less than half of the patients completed all 8 cycles of chemotherapy (16). However, this was again a very conservative approach for the chemotherapy dose by the local oncologists. Perhaps the previous assumptions for taxane-based triplet chemotherapy and clinical experience might have prejudiced the final decision on dose adjustment. Furthermore, the reduction of the dose was below 25 percent, which was considered acceptable for postoperative patients. We also presented preliminary reports on OS and DFS. Though the two-year duration was not long enough to perform any comparisons with previous reports, the present survival results were not poor for this cohort because the majority of patients (8 patients) had very late stage disease on postoperative pathological findings (ypTNM stage III).

## Conclusion

Our findings suggest that neoadjuvant chemotherapy with the FLOT regimen is safe and effective for gastric cancer patients in China. The drug dose of chemotherapy and the time interval between two chemotherapy cycles need to be adjusted for adjuvant chemotherapy. The pathological regression and survival rates were comparable but it should be reevaluated in a larger cohort. The results of this study pave the way for further studies in Asian countries.

## Data Availability

The datasets used and/or analyzed during the current study are available from the corresponding author on reasonable request.

